# Low back pain service utilization and costs: association with number of visits of chiropractic manipulation, active care, manual therapy or acupuncture. A retrospective cohort study

**DOI:** 10.1101/2022.10.28.22281664

**Authors:** David Elton, Meng Zhang

## Abstract

**Background:** For individuals with low back pain (LBP), in the absence of serious pathology clinical practice guidelines (CPG) recommend a stepped approach to management with first-line emphasis on natural history, self-care, and non-pharmaceutical therapies. For individuals with non-surgical LBP initially contacting a chiropractor (DC), physical therapist (PT), or licensed acupuncturist (LAc), the purpose of this retrospective cohort study was to examine the dose response association between the number of visits of chiropractic manipulative therapy (CMT), active care (AC), manual therapy (MT), or acupuncture, exposure to second- and third-line services, and total episode cost.

**Methods:** A national sample of individuals with a single episode of non-surgical LBP occurring in 2017-2019 was analyzed using episode of care as the unit of analysis. The primary independent variables were initial contact with either a DC, PT, or LAc, and the number of visits of CMT, AC, MT, or acupuncture. Dependent measures included rate and timing of use of 13 types of health care services and total episode cost.

**Results:** 132,199 continuously insured individuals aged 18 years and older initially contacted 21,336 different DCs, 2,734 PTs and 1,339 LAcs for a single episode of non-surgical LBP. These individuals were associated with $62,185,930 in expenditures. The most common number of visits was 1 to 3 - CMT (48.2% of episodes), AC (29.7%), MT (32.1%), and acupuncture (27.0%). For each service, 1 to 3 visits was associated with the lowest rate of exposure to second- and third-line services although rate differences between visit dose categories were generally not significant or clinically. Episode total cost and duration increased significantly with increasing number of visits. CMT was associated with lowest median total episode cost at each level of visit utilization.

**Conclusions:** For non-surgical LBP episodes initially contacting a DC, PT or LAc, 1 to 3 visits of CMT, AC, MT, or acupuncture was the most common level of utilization, associated with the lowest exposure to second- and third-line services and lowest total episode cost. Among, CMT, AC, MT, and acupuncture, CMT was associated with the lowest total episode cost at each level of utilization. A higher number of visits of CMT, AC, MT or acupuncture was associated with significantly higher total cost, without meaningful impact on exposure to second- or third-line services. Unmeasured clinical benefits may be associated with higher visit counts and warrants further study.

## Background

Low back pain (LBP) is prevalent ^1-3^ and costly ^4^ with clinical practice guidelines (CPGs) describing a stepped approach in which services are sequenced into first-, second- and third-line services.^5-7^ In the absence of red flags of serious pathology LBP CPGs emphasize, as first-line approaches, individual self-management, non-pharmaceutical and non-interventional services.^5-7^

Variation in service utilization and cost outcomes for LBP have been associated with the type ofhealth care provider (HCP) initially contacted.^8,9^ A high proportion of individuals with LBP initially seek treatment from primary care providers (PCP) and physician specialists (PS) with management emphasizing second- and third-line approaches.^10^ When initially contacted by an Individual with LBP, non-prescribing HCPs, like chiropractors (DC), physical therapists (PT), or licensed acupuncturists (LAc) are more likely to have episodes associated with an emphasis on first-line therapies with less exposure to second- and third-line services.^10^

For LBP, dose-response analyses of chiropractic manipulative therapy (CMT), active care (AC), manual therapy (MT), or acupuncture services have been conducted measuring dose using the number of visits ^11-16^ using either a specific number of visits (e.g., 0, 6, 12, 18) ^11,15^, or a range of visits (e.g., 1 to 5, 6+).^12,17^ Studies of the response to different doses of CMT, AC, MT and acupuncture have considered cost, quality-adjusted life years, disability and pain free days, and other measures of pain and function, with no clear association between dose and benefits.^11,13,17,18^ Patient willingness to pay (WTP) is an important consideration when analyzing the dose, or number of visits, of CMT, AC, MT, or acupuncture services that may be associated with co-payments or deductibles.^19^

The aim of this study was to examine the association between the number of visits of CMT, AC, MT, or acupuncture services, utilization of other healthcare services, and total cost for individuals with LBP initially contacting a DC, PT or LAc. The hypothesis was that increasing the number of visits of CMT, AC, MT, and acupuncture would be associated with increasing total episode cost and minimal impact on the rate of exposure to second- or third-line services.

## Methods

### Study design, population, setting and data sources

This is a retrospective cohort study of individuals initially contacting a DC, PT, or LAc for a single episode of non-surgical LBP during the 2017-2019 study period. An analytic database was created which included de-identified enrollment records, administrative claims data for all inpatient and outpatient services, and pharmacy prescriptions, for commercially insured enrollees. HCP demographic information and professional licensure status was incorporated from an HCP database. ZIP code level population race and ethnicity data was extracted from the US Census Bureau ^20^, socioeconomic status (SES) Area Deprivation Index (ADI) data, from the University of Wisconsin Neighborhood Atlas^® 21^ and household adjusted gross income (AGI) from the Internal Revenue Service.^22^

A review was performed to assess compliance with de-identification requirements, and as it was determined the data was de-identified or a Limited Data Set in compliance with the Health Insurance Portability and Accountability Act and customer requirements, the UnitedHealth Group Office of Human Research Affairs determined that this study was exempt from Institutional Review Board review. The study was conducted and reported based on the Strengthening the Reporting of Observational Studies in Epidemiology (STROBE) guidelines (Supplement – STROBE Checklist).^23^

The study did not include an adjustment for measurable confounders such as individual age, sex and co-morbidities ^24,25^, unknown confounders, or confounders of measurable confounders, using common yet potentially inadequate approaches such as propensity score matching ^26^ due to the inability to control for important confounders of the number of visits of CMT, AC, MT, or acupuncture for individuals with LBP initially contacting a DC, PT or LAc.. Examples of confounders not available in a retrospective cohort study include; HCP options convenient to an individual’s home, workplace or daily travel routes including public transportation if used, individual preference for specific services or type of HCP including gender or racial concordance, recommendations from family or friends and influence of HCP marketing efforts, perceived LBP severity, anticipated potential out of pocket costs and WTP, time availability to participate in multiple visits, and appointment availability within an individual’s timing expectations meeting these and other criteria.^27^ As an alternative to blurring the line between association and causality through a process that simultaneously introduces distortion and complexity into results, actual measures of individual demographic and episodic characteristics, and associations, are reported for each type of initial contact HCP, and the subsequent number of visits of each type of service.

### Cohort selection and unit of analysis

The cohort included individuals aged 18 years and older with a single complete episode of LBP commencing and ending during the calendar years 2017-2019. This timeframe was selected to follow the release of the American College of Physicians (ACP) LBP CPG ^5^ in 2017 and before the influence of the COVID-19 epidemic on care patterns in early 2020. All individuals had continuous medical and pharmacy insurance coverage during the entire study period.

Episode of care was selected as the unit of analysis. This approach has been shown to be a valid way to organize all administrative claims data associated with a condition. ^28^ The *Symmetry*^*®*^ *Episode Treatment Groups*^*®*^ *(ETG*^*®*^) and *Episode Risk Groups*^*®*^ *(ERG*^*®*^) version 9.5 methodologies and definitions were used to translate administrative claims data into episodes of care, which have been reported as a valid measurement for comparison of HCPs based on cost of care. ^29^ A previous analysis of the same underlying data indicates a low risk of misclassification bias associated with using episode of care as the unit of analysis. ^10^ Using episode of care unit of measurement has potential translational benefits in supporting the transition from fee for service to value-based episodic bundled payment arrangements.

The analysis included complete episodes defined as having at least 91-day pre- and 61-day post-episode clean periods during which no services were provided by any HCP for any LBP diagnosis. LBP episodes including a surgical procedure, or associated with diagnoses of malignant and non-malignant neoplasms, fractures and other spinal trauma, infection, congenital deformities and scoliosis, autoimmune disorders, osteoporosis, and advanced arthritis were excluded from the analysis. Individuals with multiple LBP episodes during the study period were also excluded. These exclusions were made to address a potential study limitation of individuals with more complex conditions confounding study results examining the number of visits of CMT, AC, MT, and acupuncture services.

### Variables

Data preprocessing, table generation, and initial analyses were performed using Python (*Python Language Reference, Version 3*.*7*.*5*., n.d.). To evaluate whether measures were derived from a normally distributed sample we conducted a goodness-of-fit measure using D’Agostino’s K-squared test. Non-normally distributed data are reported using the median and interquartile range (IQR).

The primary independent variables were initial contact with either a DC, PT or LAc HCP, and the number of visits of CMT, AC, MT, or acupuncture services. The study cohort was able to access DC, PT and LAc HCPs directly without a referral. For each type of HCP, the analyses focused on services provided for at least 50% of episodes. This resulted in 4 combinations of HCP and service type: DC-CMT, PT-AC, PT-MT, and LAc-acupuncture (Supplement 1).

The primary dependent variable was the rate of use of 13 types of health care services segmented into first-, second-, and third-line service categories. Service utilization reflects services provided by any type of HCP an individual saw during the complete episode of LBP. The ACP LBP CPG was used as the primary source to designate treatment interventions as first-, second-, or third line.^5^ Secondary dependent variables included the total cost of care for all reimbursed services provided by any HCP during an episode, the number of different HCP seen during an episode, and episode duration measured in days. Total episode cost included costs associated with all services provided for LBP during an episode, including those not specifically identified in the 13 categories used in the analyses. Costs for services for which an insurance claim was not submitted were not available. The episode duration was the number of days between the first and last date of service for each episode.

For each HCP and service type combination odds (OR), risk (RR) ratios, and associated 95% confidence intervals, were calculated for utilization of each service type compared to the 1 to 3 visit reference. RR were reported as this is the measure more widely understood in associational analyses and due to the tendency for ORs to exaggerate risk in situations where an outcome is relatively common. ^30^ For each HCP and service type combination, bivariate relationships were calculated comparing the 1 to 3 visit reference, with different levels of visit utilization and the percent of episodes including other services, total episode cost, the number of HCP seen during the episode, and episode duration.

## Results

The cohort consisted of 132,199 individuals, associated with 132,199 complete LBP episodes for which 21,336 DCs, 2,734 PTs and 1,339 LAcs were initially contacted. There was $62,185,930 in reimbursed health care expenditures. The pre- and post-episode clean periods were substantially longer than the *ETG*^*®*^ clean period definitions and were similar among episodes with DCs, PTs, and LAcs as the initial contact HCP. Differences were observed in attributes of individuals initially contacting DCs, PTs and LAcs. PTs were initially contacted by slightly older individuals with higher ERG® Risk Score. LAcs were initially contacted by 65% females, and individuals from zip codes with lower ADI (median 23), higher AGI (median 90,081) and lower % NHW population (median 61.1%). DCs were initially contacted by individuals from zip codes with higher ADI (median 44), lower AGI (median 67,653) and higher % NHW population (median 76.9%) (Supplement 2).

For each combination of HCP and service type, the most common number of visits was 1 to 3 (Figure 1. 48.2% of episodes in the DC-CMT analysis, 29.7% in the PT-AC analysis, 32.1% in the PT-MT analysis, and 27.0% of episodes in the LAc-acupuncture analysis had 1 to 3 visits. Within each HCP and service type combination, similar characteristics were found in individual, population, and episode attributes among visit count categories (Supplement 3).

**Figure 1.**
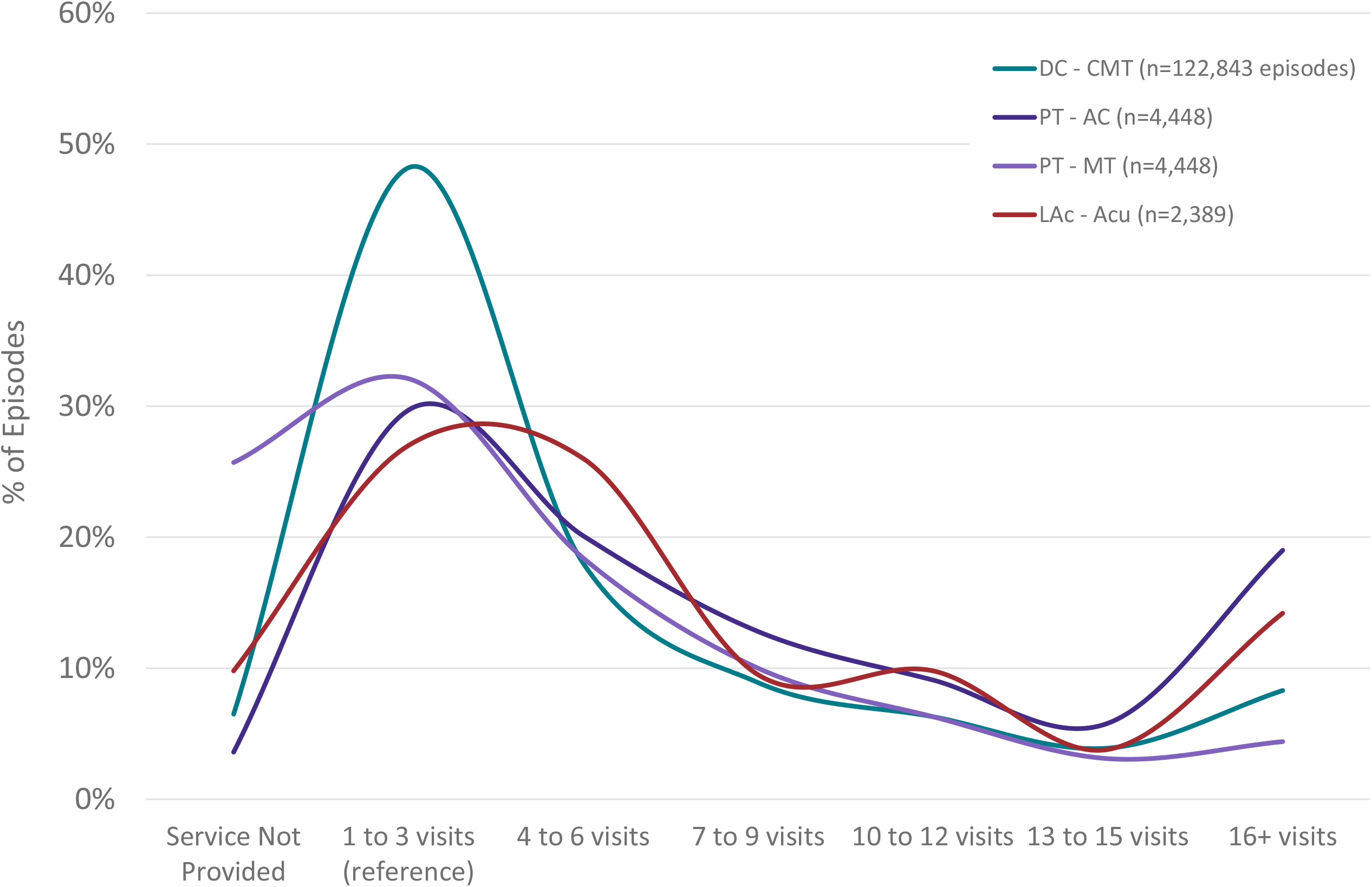
Non-surgical single episode low back pain episodedistribution by type of health care provider initially contacted and # of visits of specific type of service. DC=Doctor of Chiropractic, PT=Physical Therapist, LAc=Licensed Acupuncturist, CMT=Chiropractic Manipulative Treatment, AC=Active Care, MT=Manual Therapy, Acu=Acupuncture

**Figure 2.**
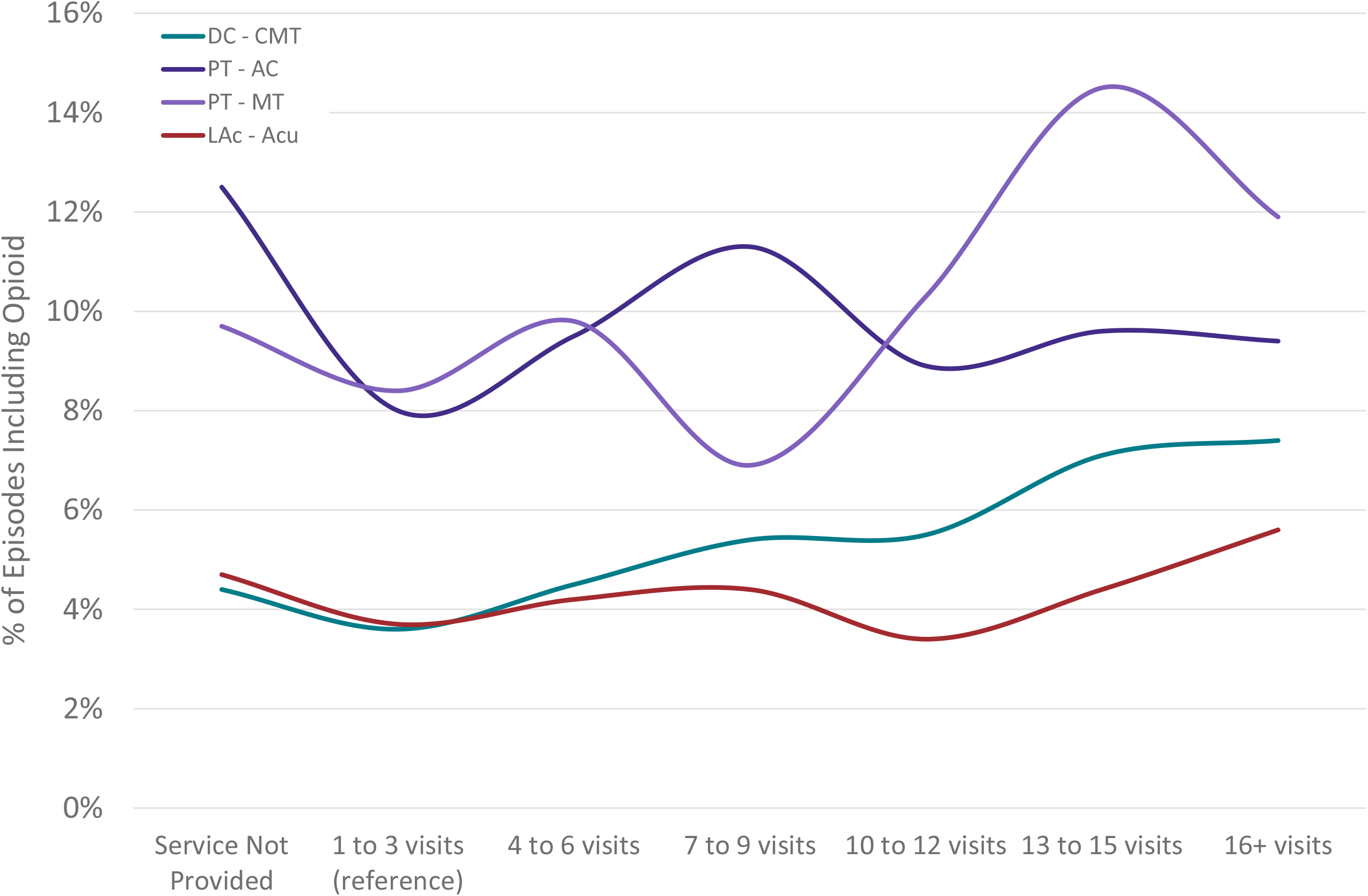
Non-surgical single episode low back pain episodes including a prescription opioid by type of health care provider initially contacted and # of visits of specific type of service. DC=Doctor of Chiropractic, PT=Physical Therapist, LAc=Licensed Acupuncturist, CMT=Chiropractic Manipulative Treatment, AC=Active Care, MT=Manual Therapy, Acu=Acupuncture

For the PT-AC, PT-MT, and LAc-acupuncture combinations, compared to the 1 to 3 visit reference category, relatively small sample sizes result in the bivariate and RR analyses revealing an absence of statistically significant or clinically meaningful differences in the exposure to second- or third-line services when AC, MT or acupuncture were not provided, or when greater than 3 visits were provided. The DC-CMT combination revealed that when CMT is not provided, or when greater than 3 visits are provided, exposure to all second- and third-line services is higher than the 1 to 3 visit reference category. (Table 1) (Table 1a). Figure 3 illustrates the RR for exposure to prescription opioids.

**Table 1.** % of single episode non-surgical low back pain episodes including specific services by type of initial contact health care provider and number of visits of select first line service

|  |  |  |  |  |  |  |  |  |  |
| --- | --- | --- | --- | --- | --- | --- | --- | --- | --- |
| % of Episodes Including |  | Service Not Provided | 1 to 3 visits (reference) | 4 to 6 visits | 7 to 9 visits | 10 to 12 visits | 13 to 15 visits | 16+ visits | Total |
| Initial Contact With Chiropractor (DC) - # of Visits of Chiropractic Manipulative Treatment (CMT) |  |  |  |  |  |  |  |  |  |
| First Line | Chiropractic Manipulation | 0.0% | 100.0% | 100.0% | 100.0% | 100.0% | 100.0% | 100.0% | 93.5% |
|  | Active Care | 44.7% | 24.2% | 32.4% | 35.8% | 37.4% | 39.7% | 45.8% | 31.3% |
|  | Manual Therapy | 22.2% | 15.5% | 19.1% | 20.2% | 20.1% | 22.4% | 24.7% | 18.3% |
|  | Acupuncture | 0.9% | 0.4% | 0.6% | 0.8% | 0.8% | 0.9% | 1.2% | 0.6% |
|  | Passive Therapy | 18.9% | 32.3% | 40.3% | 43.0% | 44.9% | 45.6% | 51.0% | 36.6% |
|  | Osteopathic Manipulation | 0.6% | 0.1% | 0.1% | 0.1% | 0.1% | 0.2% | 0.2% | 0.2% |
| Second Line | Imaging - Radiography | 38.6% | 12.5% | 20.3% | 24.6% | 28.3% | 30.9% | 36.2% | 20.3% |
|  | Rx - NSAID | 6.9% | 4.7% | 6.0% | 6.7% | 7.5% | 8.5% | 8.8% | 5.9% |
|  | Rx - Skeletal Muscle Relaxant | 5.5% | 3.4% | 4.2% | 4.8% | 4.4% | 5.8% | 6.2% | 4.2% |
|  | Imaging - MRI | 4.0% | 1.5% | 2.6% | 3.4% | 3.6% | 3.8% | 5.0% | 2.5% |
| Third Line | Rx - Opioid | 4.4% | 3.6% | 4.5% | 5.4% | 5.5% | 7.1% | 7.4% | 4.5% |
|  | Spinal Injection | 1.5% | 0.8% | 1.4% | 1.7% | 1.6% | 2.1% | 3.2% | 1.3% |
|  | Imaging - CT | 0.5% | 0.2% | 0.3% | 0.4% | 0.3% | 0.4% | 0.5% | 0.3% |
| Initial Contact With Physical Therapist (PT) - # of Visits of Active Care (AC) |  |  |  |  |  |  |  |  |  |
| First Line | Chiropractic Manipulation | 17.5% | 7.4% | 8.5% | 9.2% | 8.7% | 11.9% | 12.0% | 9.4% |
|  | Active Care | 0.0% | 100.0% | 100.0% | 100.0% | 100.0% | 100.0% | 100.0% | 96.4% |
|  | Manual Therapy | 50.6% | 59.9% | 78.2% | 81.9% | 80.4% | 85.4% | 85.7% | 74.3% |
|  | Acupuncture | 6.9% | 0.8% | 0.8% | 0.9% | 1.7% | 1.9% | 2.0% | 1.4% |
|  | Passive Therapy | 26.2% | 17.5% | 26.5% | 27.5% | 30.9% | 32.3% | 35.9% | 26.5% |
|  | Osteopathic Manipulation | 3.1% | 0.5% | 0.4% | 0.5% | 0.2% | 0.8% | 0.7% | 0.6% |
| Second Line | Imaging - Radiography | 26.2% | 20.8% | 24.4% | 26.8% | 28.0% | 28.5% | 28.1% | 25.0% |
|  | Rx - NSAID | 8.8% | 9.9% | 13.5% | 15.1% | 11.6% | 16.9% | 15.5% | 12.9% |
|  | Rx - Skeletal Muscle Relaxant | 10.0% | 8.2% | 9.9% | 10.9% | 11.6% | 11.9% | 12.7% | 10.3% |
|  | Imaging - MRI | 15.0% | 13.3% | 14.9% | 21.7% | 19.3% | 21.9% | 23.0% | 17.6% |
| Third Line | Rx - Opioid | 12.5% | 8.0% | 9.5% | 11.3% | 8.9% | 9.6% | 9.4% | 9.3% |
|  | Spinal Injection | 8.1% | 7.2% | 7.7% | 8.3% | 10.9% | 11.9% | 14.1% | 9.4% |
|  | Imaging - CT | 0.6% | 1.1% | 0.6% | 1.9% | 2.7% | 1.2% | 2.6% | 1.5% |
| Initial Contact With Physical Therapist (PT) - # of Visits of Manual Therapy (MT) |  |  |  |  |  |  |  |  |  |
| First Line | Chiropractic Manipulation | 7.5% | 8.9% | 9.3% | 9.6% | 13.2% | 12.3% | 17.5% | 9.4% |
|  | Active Care | 93.1% | 95.4% | 98.9% | 99.3% | 99.3% | 100.0% | 99.0% | 96.4% |
|  | Manual Therapy | 0.0% | 100.0% | 100.0% | 100.0% | 100.0% | 100.0% | 100.0% | 74.3% |
|  | Acupuncture | 1.3% | 0.9% | 1.5% | 1.1% | 0.7% | 5.1% | 4.6% | 1.4% |
|  | Passive Therapy | 15.5% | 22.8% | 32.4% | 36.5% | 33.1% | 44.2% | 48.5% | 26.5% |
|  | Osteopathic Manipulation | 0.3% | 0.7% | 0.2% | 1.3% | 1.1% | 1.4% | 0.5% | 0.6% |
| Second Line | Imaging - Radiography | 24.6% | 22.1% | 24.7% | 26.6% | 26.7% | 33.3% | 37.1% | 25.0% |
|  | Rx - NSAID | 11.6% | 11.1% | 13.0% | 13.4% | 16.0% | 24.6% | 19.1% | 12.9% |
|  | Rx - Skeletal Muscle Relaxant | 8.9% | 9.4% | 9.7% | 11.9% | 14.2% | 15.9% | 14.9% | 10.3% |
|  | Imaging - MRI | 15.4% | 14.9% | 16.4% | 22.8% | 22.4% | 27.5% | 30.4% | 17.6% |
| Third Line | Rx - Opioid | 9.7% | 8.4% | 9.8% | 6.9% | 10.3% | 14.5% | 11.9% | 9.3% |
|  | Spinal Injection | 7.8% | 8.5% | 9.4% | 8.9% | 11.4% | 19.6% | 16.5% | 9.4% |
|  | Imaging - CT | 1.7% | 0.8% | 1.6% | 1.1% | 2.1% | 2.9% | 4.6% | 1.5% |
| Initial Contact with Licensed Acupuncturist (LAc) - # of Visits of Acupuncture (Acu) |  |  |  |  |  |  |  |  |  |
| First Line | Chiropractic Manipulation | 24.9% | 6.2% | 5.9% | 4.4% | 7.7% | 7.8% | 10.9% | 8.7% |
|  | Active Care | 27.9% | 13.5% | 14.3% | 18.9% | 15.0% | 15.6% | 29.2% | 18.1% |
|  | Manual Therapy | 65.7% | 37.9% | 39.2% | 48.7% | 38.2% | 44.4% | 56.9% | 45.0% |
|  | Acupuncture | 0.0% | 100.0% | 100.0% | 100.0% | 100.0% | 100.0% | 100.0% | 90.2% |
|  | Passive Therapy | 53.2% | 37.3% | 41.6% | 42.5% | 34.3% | 41.1% | 50.7% | 42.2% |
|  | Osteopathic Manipulation | 0.9% | 0.2% | 0.3% | 0.0% | 0.0% | 3.3% | 0.3% | 0.4% |
| Second Line | Imaging - Radiography | 8.6% | 3.6% | 4.2% | 7.9% | 3.4% | 4.4% | 6.2% | 5.0% |
|  | Rx - NSAID | 4.7% | 4.2% | 4.0% | 6.1% | 3.4% | 3.3% | 8.6% | 4.9% |
|  | Rx - Skeletal Muscle Relaxant | 2.1% | 1.9% | 3.4% | 2.6% | 2.6% | 3.3% | 2.1% | 2.5% |
|  | Imaging - MRI | 4.7% | 3.1% | 2.3% | 3.1% | 2.6% | 3.3% | 4.7% | 3.2% |
| Third Line | Rx - Opioid | 4.7% | 3.7% | 4.2% | 4.4% | 3.4% | 4.4% | 5.6% | 4.3% |
|  | Spinal Injection | 3.0% | 1.9% | 1.1% | 0.4% | 1.3% | 3.3% | 2.9% | 1.8% |
|  | Imaging - CT | 0.0% | 0.2% | 0.3% | 0.0% | 0.4% | 0.0% | 0.6% | 0.3% |

Cells with red text denote that the effect of number of visits on service usage was found not to be significantly different from that of 1-3 visit reference (Fisher's Exact p > 0.001)
Cells with black text denote that the effect of number of visits on service usage was found to be significantly different from that of 1-3 visit reference (Fisher's Exact p < 0.001)

**Table 1a.** Risk ratio and 95% confidence interval for comparing % of single episode non-surgical low back pain episodes including specific services by type of initial contact health care provider and number of visits of select first line service to 1 to 3 visit reference

| Risk ratio and 95% confidence interval |  | Service Not Provided | 1 to 3 visits | 4 to 6 visits | 7 to 9 visits | 10 to 12 visits | 13 to 15 visits | 16+ visits |
| --- | --- | --- | --- | --- | --- | --- | --- | --- |
| Initial Contact With Chiropractor (DC) - # of Visits of Chiropractic Manipulative Treatment (CMT) |  |  |  |  |  |  |  |  |
| First Line | Chiropractic Manipulation | N/A | reference | 1.00 (1.00, 1.00) | 1.00 (1.00, 1.00) | 1.00 (1.00, 1.00) | 1.00 (1.00, 1.00) | 1.00 (1.00, 1.00) |
|  | Active Care | 1.85 (1.79, 1.90) |  | 1.34 (1.30, 1.37) | 1.48 (1.43, 1.52) | 1.54 (1.50, 1.60) | 1.64 (1.58, 1.70) | 1.89 (1.84, 1.94) |
|  | Manual Therapy | 1.43 (1.37, 1.50) |  | 1.23 (1.19, 1.28) | 1.31 (1.25, 1.36) | 1.30 (1.24, 1.36) | 1.45 (1.37, 1.53) | 1.60 (1.54, 1.66) |
|  | Acupuncture | 2.18 (1.69, 2.82) |  | 1.42 (1.15, 1.75) | 1.91 (1.50, 2.43) | 1.94 (1.48, 2.55) | 2.07 (1.49, 2.86) | 2.92 (2.36, 3.61) |
|  | Passive Therapy | 0.58 (0.56, 0.61) |  | 1.25 (1.22, 1.27) | 1.33 (1.30, 1.36) | 1.39 (1.35, 1.43) | 1.41 (1.36, 1.46) | 1.58 (1.54, 1.61) |
|  | Osteopathic Manipulation | 6.84 (4.69, 9.96) |  | 1.58 (1.03, 2.43) | 1.45 (0.82, 2.57) | 1.51 (0.79, 2.88) | 1.78 (0.85, 3.73) | 2.29 (1.40, 3.74) |
| Second Line | Imaging - Radiography | 3.10 (2.99, 3.21) |  | 1.63 (1.57, 1.68) | 1.97 (1.90, 2.05) | 2.27 (2.18, 2.37) | 2.48 (2.36, 2.60) | 2.91 (2.81, 3.01) |
|  | Rx - NSAID | 1.47 (1.35, 1.61) |  | 1.28 (1.20, 1.37) | 1.45 (1.34, 1.56) | 1.61 (1.48, 1.76) | 1.83 (1.65, 2.02) | 1.88 (1.75, 2.02) |
|  | Rx - Skeletal Muscle Relaxant | 1.62 (1.47, 1.79) |  | 1.25 (1.15, 1.34) | 1.41 (1.28, 1.55) | 1.30 (1.16, 1.45) | 1.71 (1.52, 1.94) | 1.81 (1.66, 1.98) |
|  | Imaging - MRI | 2.78 (2.45, 3.15) |  | 1.79 (1.61, 1.98) | 2.31 (2.05, 2.61) | 2.44 (2.14, 2.79) | 2.60 (2.22, 3.05) | 3.45 (3.10, 3.84) |
| Third Line | Rx-Opioid | 1.22 (1.09, 1.36) |  | 1.26 (1.17, 1.36) | 1.50 (1.37, 1.64) | 1.52 (1.38, 1.69) | 1.97 (1.77, 2.21) | 2.07 (1.91, 2.24) |
|  | Spinal Injection | 1.92 (1.57, 2.35) |  | 1.79 (1.55, 2.07) | 2.14 (1.81, 2.54) | 2.10 (1.73, 2.56) | 2.70 (2.18, 3.35) | 4.18 (3.63, 4.81) |
|  | Imaging-CT | 2.10 (1.49, 2.96) |  | 1.37 (1.04, 1.81) | 1.51 (1.07, 2.14) | 1.36 (0.90, 2.06) | 1.44 (0.87, 2.38) | 1.82 (1.31, 2.54) |
|  | Initial Contact With Physical Therapist (PT) - # of Visits of Active Care (AC) |  |  |  |  |  |  |  |
| First Line | Chiropractic Manipulation | 2.38 (1.62, 3.50) | reference | 1.16 (0.87, 1.54) | 1.24 (0.90, 1.72) | 1.18 (0.81, 1.71) | 1.62 (1.11, 2.38) | 1.63 (1.25, 2.12) |
|  | Active Care | N/A |  | 1.00 (1.00, 1.00) | 1.00 (1.00, 1.00) | 1.00 (1.00, 1.00) | 1.00 (1.00, 1.00) | 1.00 (1.00, 1.00) |
|  | Manual Therapy | 0.85 (0.72, 0.99) |  | 1.31 (1.23, 1.38) | 1.37 (1.29, 1.45) | 1.34 (1.26, 1.43) | 1.43 (1.33, 1.52) | 1.43 (1.36, 1.51) |
|  | Acupuncture | 8.24 (3.63, 18.71) |  | 0.94 (0.37, 2.42) | 1.06 (0.37, 3.02) | 2.08 (0.81, 5.32) | 2.31 (0.81, 6.58) | 2.42 (1.14, 5.13) |
|  | Passive Therapy | 1.50 (1.13, 1.99) |  | 1.52 (1.29, 1.78) | 1.57 (1.31, 1.87) | 1.77 (1.47, 2.13) | 1.84 (1.49, 2.28) | 2.05 (1.77, 2.38) |
|  | Osteopathic Manipulation | 6.87 (2.12, 22.25) |  | 0.98 (0.28, 3.48) | 1.16 (0.29, 4.63) | 0.54 (0.07, 4.51) | 1.69 (0.34, 8.33) | 1.56 (0.51, 4.83) |
| Second Line | Imaging - Radiography | 1.26 (0.95, 1.67) |  | 1.18 (1.01, 1.37) | 1.29 (1.08, 1.53) | 1.35 (1.11, 1.63) | 1.37 (1.10, 1.71) | 1.35 (1.16, 1.57) |
|  | Rx - NSAID | 0.88 (0.52, 1.49) |  | 1.36 (1.08, 1.72) | 1.52 (1.18, 1.96) | 1.17 (0.86, 1.60) | 1.70 (1.24, 2.33) | 1.56 (1.25, 1.96) |
|  | Rx - Skeletal Muscle Relaxant | 1.22 (0.74, 2.01) |  | 1.20 (0.92, 1.57) | 1.33 (0.99, 1.79) | 1.42 (1.03, 1.96) | 1.46 (1.00, 2.12) | 1.55 (1.20, 1.99) |
|  | Imaging - MRI | 1.12 (0.76, 1.67) |  | 1.12 (0.91, 1.38) | 1.62 (1.32, 2.00) | 1.45 (1.14, 1.84) | 1.64 (1.26, 2.15) | 1.72 (1.43, 2.07) |
| Third Line | Rx-Opioid | 1.57 (1.00, 2.46) |  | 1.20 (0.91, 1.57) | 1.42 (1.05, 1.90) | 1.12 (0.78, 1.61) | 1.21 (0.80, 1.83) | 1.18 (0.89, 1.55) |
|  | Spinal Injection | 1.13 (0.65, 1.97) |  | 1.07 (0.80, 1.45) | 1.15 (0.82, 1.61) | 1.51 (1.08, 2.12) | 1.66 (1.13, 2.43) | 1.96 (1.52, 2.53) |
|  | Imaging-CT | 0.55 (0.07, 4.13) |  | 0.49 (0.18, 1.35) | 1.70 (0.79, 3.68) | 2.39 (1.11, 5.17) | 1.01 (0.30, 3.48) | 2.29 (1.20, 4.39) |
| Initial Contact With Physical Therapist (PT) - # of Visits of Manual Therapy (MT) |  |  |  |  |  |  |  |  |
| First Line | Chiropractic Manipulation | 0.84 (0.65, 1.10) | reference | 1.05 (0.80, 1.37) | 1.08 (0.78, 1.50) | 1.48 (1.05, 2.09) | 1.38 (0.86, 2.23) | 1.97 (1.39, 2.79) |
|  | Active Care | 0.98 (0.96, 0.99) |  | 1.04 (1.02, 1.05) | 1.04 (1.03, 1.06) | 1.04 (1.02, 1.06) | 1.05 (1.04, 1.06) | 1.04 (1.02, 1.06) |
|  | Manual Therapy | N/A |  | 1.00 (1.00, 1.00) | 1.00 (1.00, 1.00) | 1.00 (1.00, 1.00) | 1.00 (1.00, 1.00) | 1.00 (1.00, 1.00) |
|  | Acupuncture | 1.44 (0.69, 3.01) |  | 1.61 (0.74, 3.52) | 1.23 (0.44, 3.43) | 0.78 (0.18, 3.44) | 5.57 (2.26, 13.72) | 5.09 (2.21, 11.76) |
|  | Passive Therapy | 0.68 (0.58, 0.80) |  | 1.42 (1.24, 1.63) | 1.60 (1.37, 1.87) | 1.45 (1.20, 1.76) | 1.94 (1.57, 2.40) | 2.13 (1.79, 2.53) |
|  | Osteopathic Manipulation | 0.37 (0.10, 1.36) |  | 0.35 (0.08, 1.59) | 1.92 (0.70, 5.24) | 1.52 (0.42, 5.50) | 2.07 (0.46, 9.34) | 0.74 (0.09, 5.71) |
| Second Line | Imaging - Radiography | 1.11 (0.97, 1.28) |  | 1.12 (0.96, 1.31) | 1.21 (1.01, 1.45) | 1.21 (0.97, 1.50) | 1.51 (1.17, 1.95) | 1.68 (1.37, 2.07) |
|  | Rx - NSAID | 1.04 (0.84, 1.30) |  | 1.16 (0.92, 1.47) | 1.20 (0.91, 1.59) | 1.44 (1.06, 1.95) | 2.21 (1.60, 3.07) | 1.71 (1.24, 2.37) |
|  | Rx - Skeletal Muscle Relaxant | 0.95 (0.74, 1.21) |  | 1.03 (0.79, 1.34) | 1.26 (0.94, 1.70) | 1.52 (1.09, 2.11) | 1.70 (1.12, 2.57) | 1.59 (1.10, 2.31) |
|  | Imaging - MRI | 1.03 (0.86, 1.24) |  | 1.10 (0.90, 1.34) | 1.53 (1.24, 1.89) | 1.50 (1.17, 1.93) | 1.84 (1.37, 2.48) | 2.04 (1.59, 2.61) |
| Third Line | Rx-Opioid | 1.15 (0.90, 1.48) |  | 1.16 (0.89, 1.52) | 0.82 (0.56, 1.21) | 1.23 (0.84, 1.80) | 1.72 (1.11, 2.68) | 1.41 (0.93, 2.15) |
|  | Spinal Injection | 0.92 (0.71, 1.19) |  | 1.11 (0.85, 1.46) | 1.06 (0.75, 1.48) | 1.34 (0.93, 1.94) | 2.31 (1.58, 3.37) | 1.95 (1.36, 2.79) |
|  | Imaging-CT | 2.27 (1.09, 4.71) |  | 2.06 (0.93, 4.59) | 1.45 (0.51, 4.15) | 2.77 (1.03, 7.43) | 3.76 (1.21, 11.65) | 6.02 (2.53, 14.34) |
| Initial Contact with Licensed Acupuncturist (LAc) - # of Visits of Acupuncture (Acu) |  |  |  |  |  |  |  |  |
| First Line | Chiropractic Manipulation | 4.01 (2.76, 5.82) | reference | 0.96 (0.62, 1.48) | 0.71 (0.36, 1.39) | 1.24 (0.73, 2.13) | 1.25 (0.58, 2.71) | 1.76 (1.15, 2.69) |
|  | Active Care | 2.07 (1.55, 2.74) |  | 1.06 (0.81, 1.39) | 1.40 (1.00, 1.95) | 1.11 (0.77, 1.60) | 1.15 (0.68, 1.94) | 2.16 (1.67, 2.79) |
|  | Manual Therapy | 1.73 (1.51, 1.98) |  | 1.04 (0.90, 1.19) | 1.28 (1.09, 1.52) | 1.01 (0.83, 1.22) | 1.17 (0.91, 1.51) | 1.50 (1.31, 1.72) |
|  | Acupuncture | N/A |  | 1.00 (1.00, 1.00) | 1.00 (1.00, 1.00) | 1.00 (1.00, 1.00) | 1.00 (1.00, 1.00) | 1.00 (1.00, 1.00) |
|  | Passive Therapy | 1.43 (1.22, 1.67) |  | 1.12 (0.97, 1.28) | 1.14 (0.95, 1.37) | 0.92 (0.75, 1.13) | 1.10 (0.84, 1.44) | 1.36 (1.18, 1.57) |
|  | Osteopathic Manipulation | 5.53 (0.50, 60.68) |  | 2.07 (0.19, 22.78) | N/A | N/A | 1.47 (2.26, 204.16) | 1.90 (0.12, 30.28) |
| Second Line | Imaging - Radiography | 2.40 (1.35, 4.29) |  | 1.17 (0.68, 2.03) | 2.21 (1.22, 4.02) | 0.96 (0.44, 2.12) | 1.24 (0.44, 3.52) | 1.73 (0.97, 3.09) |
|  | Rx - NSAID | 1.13 (0.57, 2.23) |  | 0.96 (0.56, 1.63) | 1.46 (0.78, 2.74) | 0.82 (0.38, 1.78) | 0.80 (0.25, 2.57) | 2.04 (1.23, 3.39) |
|  | Rx - Skeletal Muscle Relaxant | 1.15 (0.41, 3.23) |  | 1.81 (0.90, 3.65) | 1.41 (0.54, 3.72) | 1.38 (0.52, 3.64) | 1.79 (0.51, 6.22) | 1.11 (0.44, 2.79) |
|  | Imaging - MRI | 1.52 (0.74, 3.12) |  | 0.72 (0.37, 1.42) | 0.99 (0.42, 2.31) | 0.83 (0.34, 2.04) | 1.07 (0.33, 3.54) | 1.52 (0.80, 2.89) |
| Third Line | Rx-Opioid | 1.27 (0.63, 2.55) |  | 1.12 (0.65, 1.93) | 1.18 (0.57, 2.42) | 0.92 (0.42, 2.02) | 1.19 (0.42, 3.36) | 1.50 (0.84, 2.71) |
|  | Spinal Injection | 1.61 (0.64, 4.05) |  | 0.60 (0.24, 1.52) | 0.24 (0.03, 1.80) | 0.69 (0.20, 2.43) | 1.79 (0.51, 6.22) | 1.58 (0.69, 3.63) |
|  | Imaging-CT | N/A |  | 2.07 (0.19, 22.78) | N/A | 2.76 (0.17, 44.01) | N/A | 3.80 (0.35, 41.75) |

Cells with red text denote 95% confidence interval includes 1 Cells with black text denote 95% confidence interval does not include 1

**Figure 3.**
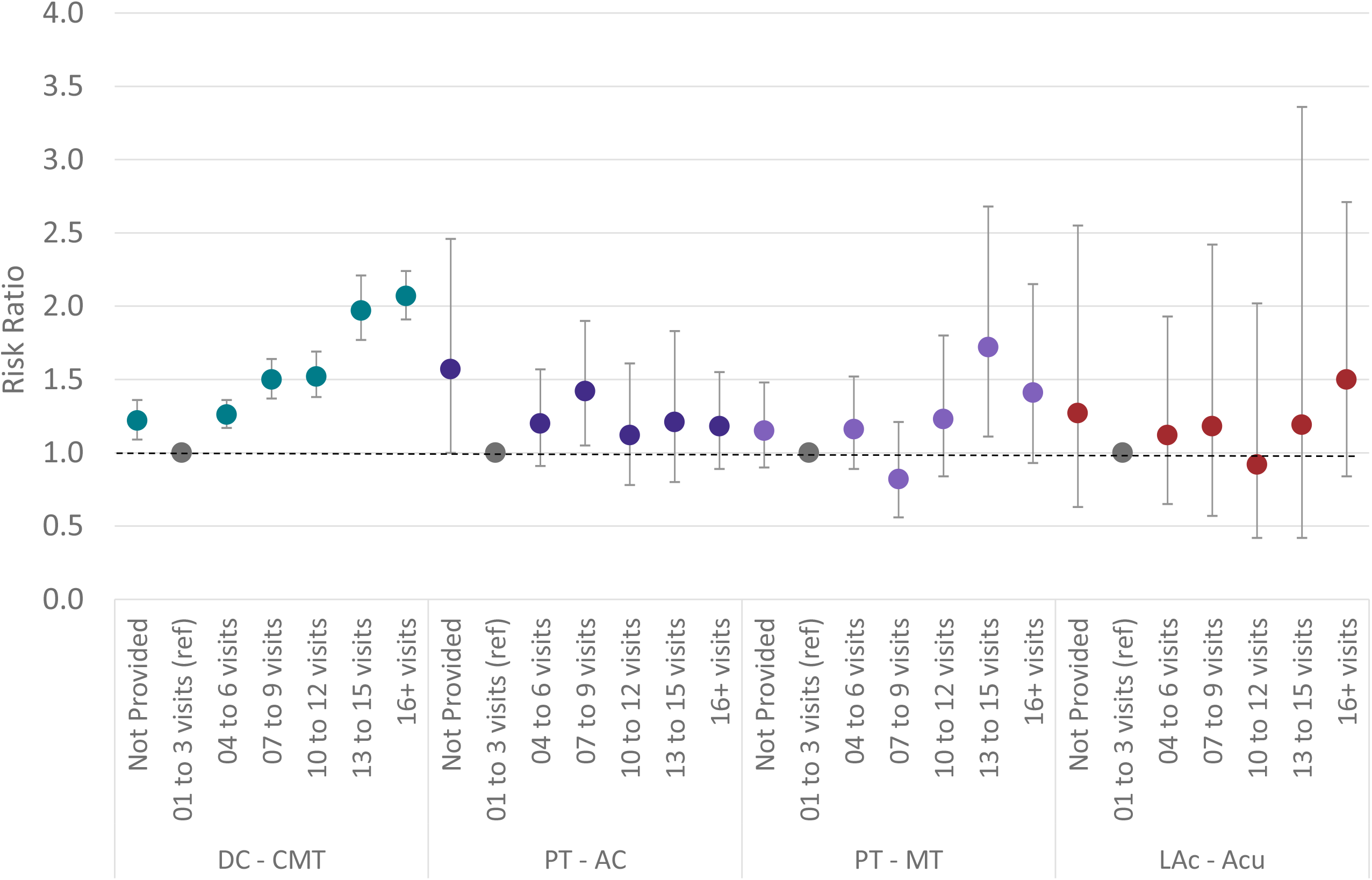
Non-surgical single episode low back pain risk ratio and 95% confidence interval for opioid exposure compared to the 1 to 3 visit reference by type of health care provider initially contacted and # of visits of specific type of service. DC=Doctor of Chiropractic, PT=Physical Therapist, LAc=Licensed Acupuncturist, CMT=Chiropractic Manipulative Treatment, AC=Active Care, MT=Manual Therapy, Acu=Acupuncture

For the DC-CMT, PT-AC, PT-MT, and LAc-acupuncture combinations, significant increases in total episode cost and episode duration were associated with an increasing number of visits. Within each visit count category total episode cost was lowest for the DC-CMT combination and the DC-CMT combination was also associated with the lowest overall median total episode cost ($194, Q1 88 Q3 457) (Table 2 (Figure 4). As the number of visits increased, the number of different HCPs seen during an episode increased for the PT-AC and PT-MT combinations and was unchanged for the DC-CMT and LAc-acupuncture combinations.

**Table 2.**
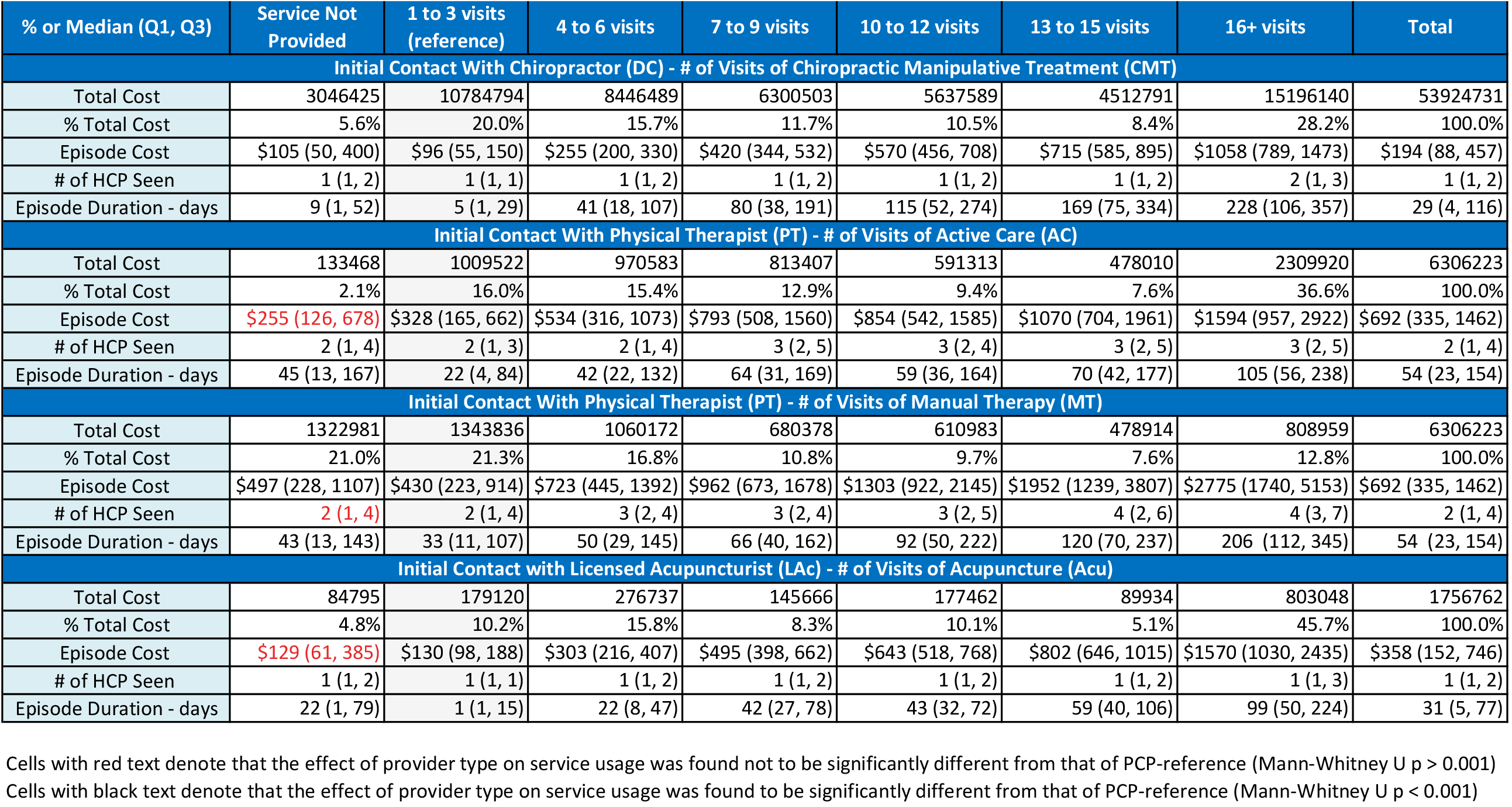
Single episode non-surgical low back pain episode characteristics by type of initial contact health care provider (HCP) and number of visits of select services

| % or Median (Q1, Q3) | Service Not Provided | 1 to 3 visits (reference) | 4 to 6 visits | 7 to 9 visits | 10 to 12 visits | 13 to 15 visits | 16+ visits | Total |
| --- | --- | --- | --- | --- | --- | --- | --- | --- |
| **Initial Contact With Chiropractor (DC) - # of Visits of Chiropractic Manipulative Treatment (CMT)** |  |  |  |  |  |  |  |  |
| Total Cost | 3046425 | 10784794 | 8446489 | 6300503 | 5637589 | 4512791 | 15196140 | 53924731 |
| % Total Cost | 5.6% | 20.0% | 15.7% | 11.7% | 10.5% | 8.4% | 28.2% | 100.0% |
| Episode Cost | \$105 (50, 400) | \$96 (55, 150) | \$255 (200, 330) | \$420 (344, 532) | \$570 (456, 708) | \$715 (585, 895) | \$1058 (789, 1473) | \$194 (88, 457) |
| # of HCP Seen | 1 (1, 2) | 1 (1, 1) | 1 (1, 2) | 1 (1, 2) | 1 (1, 2) | 1 (1, 2) | 2 (1, 3) | 1 (1, 2) |
| Episode Duration - days | 9 (1, 52) | 5 (1, 29) | 41 (18, 107) | 80 (38, 191) | 115 (52, 274) | 169 (75, 334) | 228 (106, 357) | 29 (4, 116) |
| **Initial Contact With Physical Therapist (PT) - # of Visits of Active Care (AC)** |  |  |  |  |  |  |  |  |
| Total Cost | 133468 | 1009522 | 970583 | 813407 | 591313 | 478010 | 2309920 | 6306223 |
| % Total Cost | 2.1% | 16.0% | 15.4% | 12.9% | 9.4% | 7.6% | 36.6% | 100.0% |
| Episode Cost | \$255 (126, 678) | \$328 (165, 662) | \$534 (316, 1073) | \$793 (508, 1560) | \$854 (542, 1585) | \$1070 (704, 1961) | \$1594 (957, 2922) | \$692 (335, 1462) |
| # of HCP Seen | 2 (1, 4) | 2 (1, 3) | 2 (1, 4) | 3 (2, 5) | 3 (2, 4) | 3 (2, 5) | 3 (2, 5) | 2 (1, 4) |
| Episode Duration - days | 45 (13, 167) | 22 (4, 84) | 42 (22, 132) | 64 (31, 169) | 59 (36, 164) | 70 (42, 177) | 105 (56, 238) | 54 (23, 154) |
| **Initial Contact With Physical Therapist (PT) - # of Visits of Manual Therapy (MT)** |  |  |  |  |  |  |  |  |
| Total Cost | 1322981 | 1343836 | 1060172 | 680378 | 610983 | 478914 | 808959 | 6306223 |
| % Total Cost | 21.0% | 21.3% | 16.8% | 10.8% | 9.7% | 7.6% | 12.8% | 100.0% |
| Episode Cost | \$497 (228, 1107) | \$430 (223, 914) | \$723 (445, 1392) | \$962 (673, 1678) | \$1303 (922, 2145) | \$1952 (1239, 3807) | \$2775 (1740, 5153) | \$692 (335, 1462) |
| # of HCP Seen | 2 (1, 4) | 2 (1, 4) | 3 (2, 4) | 3 (2, 4) | 3 (2, 5) | 4 (2, 6) | 4 (3, 7) | 2 (1, 4) |
| Episode Duration - days | 43 (13, 143) | 33 (11, 107) | 50 (29, 145) | 66 (40, 162) | 92 (50, 222) | 120 (70, 237) | 206 (112, 345) | 54 (23, 154) |
| **Initial Contact with Licensed Acupuncturist (LAc) - # of Visits of Acupuncture (Acu)** |  |  |  |  |  |  |  |  |
| Total Cost | 84795 | 179120 | 276737 | 145666 | 177462 | 89934 | 803048 | 1756762 |
| % Total Cost | 4.8% | 10.2% | 15.8% | 8.3% | 10.1% | 5.1% | 45.7% | 100.0% |
| Episode Cost | \$129 (61, 385) | \$130 (98, 188) | \$303 (216, 407) | \$495 (398, 662) | \$643 (518, 768) | \$802 (646, 1015) | \$1570 (1030, 2435) | \$358 (152, 746) |
| # of HCP Seen | 1 (1, 2) | 1 (1, 1) | 1 (1, 2) | 1 (1, 2) | 1 (1, 2) | 1 (1, 2) | 1 (1, 3) | 1 (1, 2) |
| Episode Duration - days | 22 (1, 79) | 1 (1, 15) | 22 (8, 47) | 42 (27, 78) | 43 (32, 72) | 59 (40, 106) | 99 (50, 224) | 31 (5, 77) |

Cells with red text denote that the effect of provider type on service usage was found not to be significantly different from that of PCP-reference (Mann-Whitney U p > 0.001)
Cells with black text denote that the effect of provider type on service usage was found to be significantly different from that of PCP-reference (Mann-Whitney U p < 0.001)

**Figure 4.**
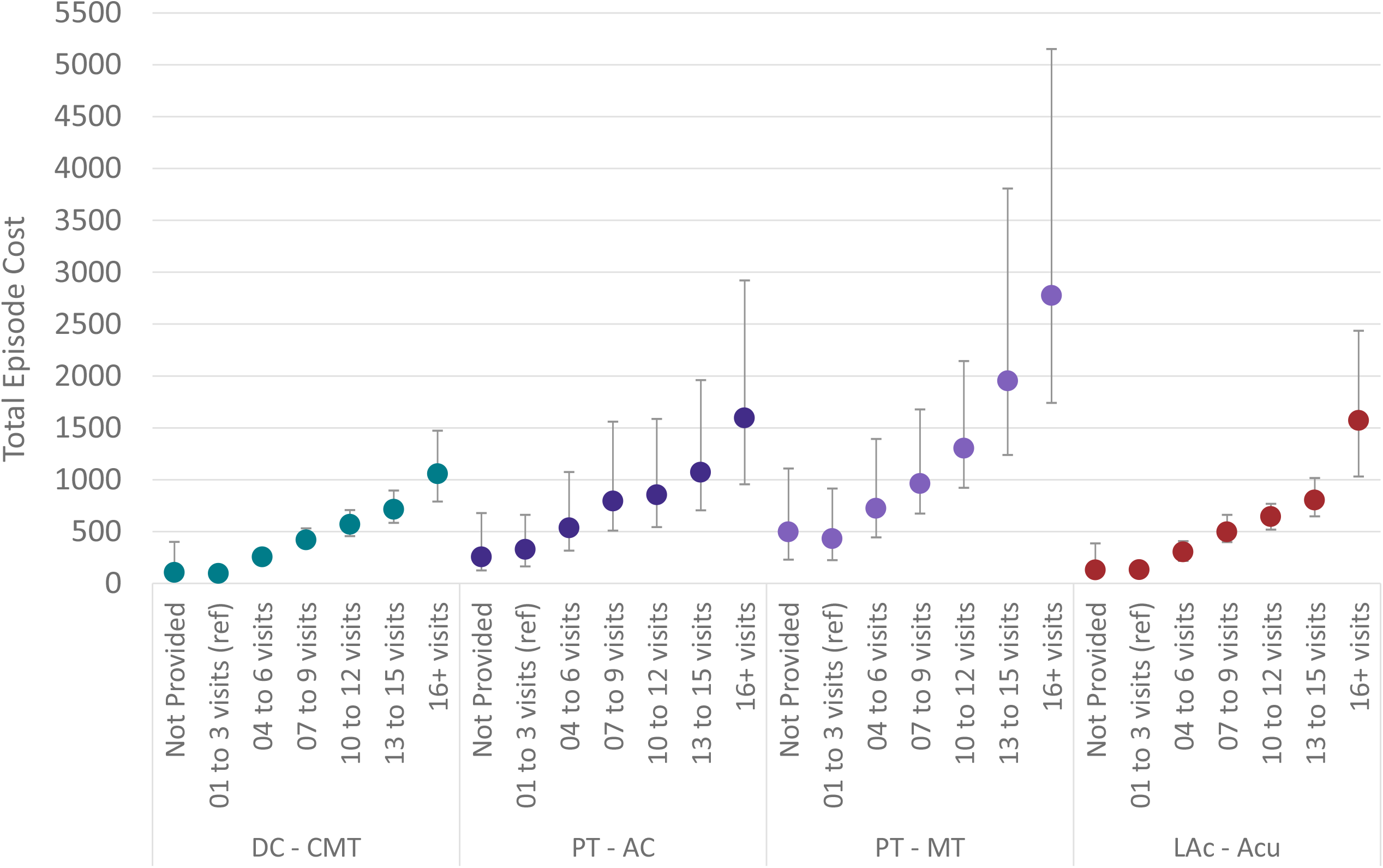
Non-surgical single episode low back pain median and interquartile range (Q1,Q3) total episode cost by type of health care provider initially contacted and # of visits of specific type of service. DC=Doctor of Chiropractic, PT=Physical Therapist, LAc=Licensed Acupuncturist, CMT=Chiropractic Manipulative Treatment, AC=Active Care, MT=Manual Therapy, Acu=Acupuncture

## Discussion

For individuals with non-surgical LBP initially contacting a DC, PT or LAc, this study provides a comprehensive analysis of the relationship between the number of visits of CMT, AC, MT, and acupuncture, utilization of other healthcare services, and total episode cost. The most common level of utilization of CMT, AC, MT, and acupuncture was 1 to 3 visits, with this level of utilization associated with the lowest rate of exposure to second- and third-line services, and lowest total episode cost. At all levels of utilization CMT was associated with lowest median total episode cost compared to AC, MT, and acupuncture. The study was not able to evaluate important considerations such as patient preference or WTP for number of visits of a service, or clinical benefits associated with different numbers of visits of CMT, AC, MT, and acupuncture.

This study has several limitations including its retrospective design and those associated with the use of administrative databases. The cohort had continuous highly uniform commercial insurance coverage and the processing of administrative claims data included extensive quality and actuarial control measures, nonetheless, data errors, variability in benefit plan design, variability in enrollee cost-sharing responsibility, and missing information were potential sources of confounding or bias. Although the commercial insurer HCP database is under continual validation it may have included errors or missing information. Summarizing total episode cost has potential limitations associated with insurance coverage, nature of network participation, and alternative reimbursement models. While individuals from all 50 states and most US territories were included, providing a measure of generalizability, the cohort did not describe a U.S. representative sample.

Another important limitation was the risk of confounding and bias associated with the limited ability to control for individual preference for type of initial contact HCP, individual expectations, or requests for specific health care services, WTP for different doses of services, and potentially meaningful differences in clinical complexity of individuals seeking and receiving different levels of utilization of CMT, AC, MT, and acupuncture. This limitation was partially addressed by focusing on episodes where a DC, PT or LAc was initially contacted by an individual with LBP and by narrowing the study population with several exclusions. The study excluded LBP associated with significant pathology, individuals with multiple episodes of LBP, and episodes involving a surgical procedure. The absence of baseline and sequential patient reported outcome data prevented an analysis of change in patient functional status associated with different numbers of visits of CMT, AC, MT, and acupuncture.

This study corroborates and expands upon previous studies exploring dose response of CMT, AC, MT, and acupuncture for the treatment of LBP. When initially contacted by an individual with LBP, DCs, PTs and LAcs are associated with greater probability of timely incorporation of first-line services than primary care, physician specialist, and emergency medicine/urgent care HCPs.^10^ This study expands on this to demonstrate this guideline concordance benefit is associated with as few as 1 to 3 visits of CMT, AC, MT, and acupuncture services. A recent systematic review of the dose-response relationship of stabilization exercises for chronic non-specific low back pain found low quality evidence supporting 3 to 5 visits per week for a duration of 6 weeks to be feasible and effective.^13^ For an episode of non-surgical LBP this current study was not able to demonstrate that this level of utilization of AC (18 to 30 visits) was either common or associated with second- or third-line service avoidance benefits. A small, practice-based randomized controlled trial comparing dose response of light massage with CMT for chronic LBP found small but statistically and clinically insignificant benefits to 12 visits of CMT, with benefits of all dose levels observed in the first 6 weeks of the 52-week measurement period.^15^ This current study found second and third line service avoidance benefits of CMT were associated with 1 to 3 visits.

## Conclusions

For individuals initially contacting a DC, PT or LAc for a single episode of non-surgical LBP, 1 to 3 visits of CMT, AC, MT, or acupuncture was associated with guideline concordant avoidance of second- and third-line services and low total episode cost. Increasing the number of visits was not associated with additional second- and third-line service avoidance benefits, however, it was associated with significant increases in total episode cost. Individual preferences and WTP may have contributed to a higher number of visits, and there may have been unmeasured clinical benefits associated with a higher number of visits.

## Supporting information

Supplement - State Summary

Supplement - STROBE Checklist

Supplement - Episode Distribution

Supplement - Cohort Summary

Supplement - Cohort Detail

## Data Availability

All data produced in the present work are contained in the manuscript

## List of Abbreviations

LBP: Low back pain
US: United States
CPG: Clinical practice guideline
DC: Doctor of Chiropractic
PT: Physical Therapist
HCP: Health care provider
ADI: Area Deprivation Index
WTP: Willingness to pay
STROBE: Strengthening the Reporting of Observational Studies in Epidemiology
ETG^*®*^: Episode Treatment Group^*®*^
ERG^*®*^: Episode Risk Group^*®*^
ACP: American College of Physicians
RR: Risk ratio
OR: Odds ratio
SD: Standard deviation
CMT: Chiropractic manipulative treatment
AC: Active care
MT: Manual therapy

