## Supplement - State Summary for "Low back pain service utilization and costs: association with number of visits of chiropractic manipulation, active care, manual therapy or acupuncture. A retrospective cohort study"

### Supplement - Episode count by State of individual's home address

| State | Total # | Total % | DC | PT | LAc |
|---|---|---|---|---|---|
| Total | 132199 | 100.0% | 125279 | 4490 | 2430 |
| TX | 15375 | 11.6% | 15001 | 232 | 142 |
| FL | 10258 | 7.8% | 9856 | 254 | 148 |
| MO | 8300 | 6.3% | 8241 | 53 | 6 |
| WI | 7438 | 5.6% | 7202 | 226 | 10 |
| MN | 6734 | 5.1% | 6373 | 301 | 60 |
| OH | 6628 | 5.0% | 6475 | 134 | 19 |
| IL | 6015 | 4.5% | 5716 | 254 | 45 |
| CA | 5880 | 4.4% | 4963 | 260 | 657 |
| CO | 4452 | 3.4% | 4043 | 310 | 99 |
| NC | 3925 | 3.0% | 3760 | 155 | 10 |
| AZ | 3671 | 2.8% | 3536 | 119 | 16 |
| IA | 3609 | 2.7% | 3565 | 43 | 1 |
| GA | 3402 | 2.6% | 3260 | 124 | 18 |
| VA | 3296 | 2.5% | 3058 | 180 | 58 |
| WA | 3287 | 2.5% | 2943 | 116 | 228 |
| NY | 3265 | 2.5% | 2616 | 432 | 217 |
| NE | 3027 | 2.3% | 2987 | 39 | 1 |
| IN | 2765 | 2.1% | 2663 | 101 | 1 |
| MD | 2621 | 2.0% | 2233 | 194 | 194 |
| PA | 2487 | 1.9% | 2380 | 91 | 16 |
| TN | 2059 | 1.6% | 2011 | 47 | 1 |
| OR | 2011 | 1.5% | 1714 | 64 | 233 |
| LA | 1695 | 1.3% | 1659 | 35 | 1 |
| OK | 1674 | 1.3% | 1653 | 20 | 1 |
| KS | 1643 | 1.2% | 1603 | 37 | 3 |
| NJ | 1623 | 1.2% | 1493 | 79 | 51 |
| MI | 1467 | 1.1% | 1421 | 43 | 3 |
| UT | 1431 | 1.1% | 1388 | 42 | 1 |
| KY | 1292 | 1.0% | 1241 | 48 | 3 |
| AR | 1199 | 0.9% | 1165 | 33 | 1 |
| MA | 1149 | 0.9% | 1080 | 52 | 17 |
| CT | 1006 | 0.8% | 927 | 67 | 12 |
| RI | 829 | 0.6% | 742 | 60 | 27 |
| SC | 825 | 0.6% | 788 | 36 | 1 |
| MS | 772 | 0.6% | 751 | 19 | 2 |
| NV | 623 | 0.5% | 587 | 27 | 9 |
| ND | 502 | 0.4% | 499 | 3 | 0 |
| NM | 492 | 0.4% | 419 | 4 | 69 |
| AL | 421 | 0.3% | 401 | 20 | 0 |
| DC | 386 | 0.3% | 324 | 42 | 20 |
| SD | 281 | 0.2% | 277 | 4 | 0 |
| WV | 250 | 0.2% | 239 | 10 | 1 |
| ID | 236 | 0.2% | 228 | 4 | 4 |
| NH | 208 | 0.2% | 194 | 12 | 2 |
| ME | 171 | 0.1% | 160 | 8 | 3 |
| WY | 155 | 0.1% | 151 | 4 | 0 |
| VI | 143 | 0.1% | 136 | 7 | 0 |
| MT | 127 | 0.1% | 120 | 7 | 0 |
| DE | 117 | 0.1% | 111 | 4 | 2 |
| VT | 26 | 0.0% | 24 | 2 | |
| AK | 12 | 0.0% | 11 | 0 | 1 |
| HI | 10 | 0.0% | 8 | 1 | 1 |
| PR | 3 | 0.0% | 3 | 0 | 0 |
| Unknown | 926 | 0.7% | 880 | 31 | 15 |
