## Supplement - Episode Distribution for "Low back pain service utilization and costs: association with number of visits of chiropractic manipulation, active care, manual therapy or acupuncture. A retrospective cohort study"

| Supplement 1 - Single episode non-surgical low back pain episode distribution by type of initial contact health care provider and number of visits of service | | | | | | | | | |
|---|---|---|---|---|---|---|---|---|---|
| HCP Type | Service Type | # of Visits of Service | | | | | | | |
| | | Service Not Provided | 1 to 3 visits (reference) | 4 to 6 visits | 7 to 9 visits | 10 to 12 visits | 13 to 15 visits | 16+ visits | Total |
| Episode Count | | | | | | | | | |
| DC | CMT | 8042 | 59200 | 22062 | 10906 | 7715 | 4753 | 10165 | 122843 |
| | AC | 84440 | 21845 | 6901 | 3290 | 2051 | 1309 | 3007 | 122843 |
| | MT | 100363 | 13928 | 3840 | 1812 | 1168 | 660 | 1072 | 122843 |
| | Acu | 122059 | 431 | 166 | 57 | 52 | 15 | 63 | 122843 |
| PT | AC | 160 | 1319 | 893 | 568 | 404 | 260 | 844 | 4448 |
| | MT | 1144 | 1427 | 817 | 447 | 281 | 138 | 194 | 4448 |
| | CMT | 4028 | 178 | 83 | 54 | 35 | 20 | 50 | 4448 |
| | Acu | 4385 | 18 | 16 | 10 | 5 | 2 | 12 | 4448 |
| LAc | Acu | 233 | 644 | 622 | 228 | 233 | 90 | 339 | 2389 |
| | MT | 1315 | 630 | 209 | 90 | 66 | 21 | 58 | 2389 |
| | AC | 1957 | 230 | 72 | 38 | 32 | 18 | 42 | 2389 |
| | CMT | 2182 | 101 | 37 | 23 | 18 | 5 | 23 | 2389 |
| % of Episodes | | | | | | | | | |
| DC | CMT | 6.5% | 48.2% | 18.0% | 8.9% | 6.3% | 3.9% | 8.3% | 100.0% |
| | AC | 68.7% | 17.8% | 5.6% | 2.7% | 1.7% | 1.1% | 2.4% | 100.0% |
| | MT | 81.7% | 11.3% | 3.1% | 1.5% | 1.0% | 0.5% | 0.9% | 100.0% |
| | Acu | 99.4% | 0.4% | 0.1% | 0.0% | 0.0% | 0.0% | 0.1% | 100.0% |
| PT | AC | 3.6% | 29.7% | 20.1% | 12.8% | 9.1% | 5.8% | 19.0% | 100.0% |
| | MT | 25.7% | 32.1% | 18.4% | 10.0% | 6.3% | 3.1% | 4.4% | 100.0% |
| | CMT | 90.6% | 4.0% | 1.9% | 1.2% | 0.8% | 0.4% | 1.1% | 100.0% |
| | Acu | 98.6% | 0.4% | 0.4% | 0.2% | 0.1% | 0.0% | 0.3% | 100.0% |
| LAc | Acu | 9.8% | 27.0% | 26.0% | 9.5% | 9.8% | 3.8% | 14.2% | 100.0% |
| | MT | 55.0% | 26.4% | 8.7% | 3.8% | 2.8% | 0.9% | 2.4% | 100.0% |
| | AC | 81.9% | 9.6% | 3.0% | 1.6% | 1.3% | 0.8% | 1.8% | 100.0% |
| | CMT | 91.3% | 4.2% | 1.5% | 1.0% | 0.8% | 0.2% | 1.0% | 100.0% |

DC=Doctor of Chiropractic, PT=Physical Therapist, LAc=Licensed Acupuncturist, CMT=Chiropractic Manipulative Treatment, AC=Active Care, MT=Manual Therapy, Acu=Acupuncture
