## Supplement - Cohort Summary for "Low back pain service utilization and costs: association with number of visits of chiropractic manipulation, active care, manual therapy or acupuncture. A retrospective cohort study"

**Supplement 2 - Non-surgical single episode low back pain cohort characteristics for individuals initially contacting a Chiropractor (DC), Physical Therapist (PT), or Licensed Acupuncturists (LAc)**

| | DC | PT | LAc |
|---|---|---|---|
| # Unique Health Care Providers (HCP) | 21336 | 2734 | 1339 |
| Episodes | 125279 | 4490 | 2430 |
| Individuals | 125279 | 4490 | 2430 |
| Total Cost | 54114697 | 6311324 | 1759909 |
| Individuals - % or (Median (Q1,Q3)) | | | |
| % Female | 51.4% | 58.9% | 65.4% |
| Age | 40 (31, 50) | 45 (34, 55) | 40 (33, 50) |
| ERG® Risk Score | 1.0 (0.4, 2.0) | 1.7 (0.8, 3.3) | 1.0 (0.5, 2.4) |
| Individual Home Address 5 Digit Zip Code Population Attributes - (Median (Q1,Q3)) | | | |
| % Non-Hispanic White (NHW) | 76.9% (59.9%, 88.1%) | 73.0% (54.6%, 84.3%) | 61.1% (40.4%, 76.5%) |
| Area Deprivation Index (ADI) | 44 (28, 60) | 34 (19, 52) | 23 (12, 36) |
| Household Adjusted Gross Income (AGI) | 67653 (53337, 93055) | 77685 (56821, 114490) | 90081 (64357, 132403) |
| DC per 1000 Population | 0.27 (0.12, 0.50) | 0.27 (0.12, 0.48) | 0.27 (0.13, 0.51) |
| PT per 1000 Population | 0.19 (0.05, 0.45) | 0.25 (0.09, 0.58) | 0.25 (0.09, 0.57) |
| LAc per 1000 Population | 0.00 (0.00, 0.03) | 0.00 (0.00, 0.06) | 0.06 (0.00, 0.19) |
| Episode Attributes - % or (Median (Q1,Q3)(Minimum)) | | | |
| % Without CMT, AC, MT and Acu | 1.9% | 0.9% | 1.7% |
| Episode Cost | 185 (80, 450) | 682 (327, 1451) | 352 (146, 733) |
| # of Different HCP Seen | 1 (1, 2) | 2 (1, 4) | 1 (1, 2) |
| Episode Duration - Days | 29 (4, 113) (1) | 53 (22, 152) (1) | 30 (4, 74) (1) |
| Clean Period Before Episode - Days | 672 (459, 879) (91) | 629 (444, 842) (91) | 681 (458, 891) (91) |
| Clean Period After Episode - Days | 397 (236, 627) (61) | 406 (246, 643) (61) | 418 (235, 655) (61) |

CMT=Chiropractic Manipulative Treatment, AC=Active Care, MT=Manual Therapy, Acu=Acupuncture
