## Supplement - Cohort Detail for "Low back pain service utilization and costs: association with number of visits of chiropractic manipulation, active care, manual therapy or acupuncture. A retrospective cohort study"

| Supplement 3 - Single episode non-surgical low back pain cohort characteristics by type of initial contact health care provider and number of visits of select first line service | | | | | | | | |
|---|---|---|---|---|---|---|---|---|
| % or Median (Q1, Q3) | Service Not Provided | 1 to 3 visits (reference) | 4 to 6 visits | 7 to 9 visits | 10 to 12 visits | 13 to 15 visits | 16+ visits | Total |
| **Initial Contact With Chiropractor (DC) - # of Visits of Chiropractic Manipulative Treatment (CMT)** | | | | | | | | |
| # of DCs | 564 | 16231 | 10817 | 6982 | 5400 | 3631 | 5926 | 21215 |
| % of DCs | 2.7% | 76.5% | 51.0% | 32.9% | 25.5% | 17.1% | 27.9% | 100.0% |
| Episodes/Individuals | 8042 | 59200 | 22062 | 10906 | 7715 | 4753 | 10165 | 122843 |
| % of Episodes/Individuals | 6.5% | 48.2% | 18.0% | 8.9% | 6.3% | 3.9% | 8.3% | 100.0% |
| Individuals - % Female | 53.3% | 48.7% | 51.7% | 54.0% | 54.6% | 55.1% | 57.8% | 51.4% |
| Individuals - Age | 39 (30, 49) | 38 (29, 49) | 40 (31, 51) | 41 (32, 51) | 41 (32, 52) | 42 (32, 52) | 42 (33, 53) | 40 (31, 50) |
| Individuals - ERG® Risk Score | 1.0 (0.5, 2.1) | 0.9 (0.4, 1.8) | 1.0 (0.5, 2.1) | 1.1 (0.5, 2.2) | 1.1 (0.5, 2.2) | 1.1 (0.6, 2.3) | 1.2 (0.6, 2.5) | 1.0 (0.4, 2.0) |
| Zip Code - % non-Hispanic White | 67% (49, 82) | 78% (61, 89) | 77% (61, 88%) | 78% (61, 88) | 78% (62, 89) | 78% (62, 88) | 77% (61, 88) | 77% (60, 88) |
| Zip Code - Area Deprivation Index | 43 (28, 60) | 45 (29, 61) | 43 (27, 59) | 43 (27, 59) | 42 (27, 58) | 43 (27, 59) | 41 (26, 57) | 44 (28, 60) |
| Zip Code - DCs per 1000 | 0.22 (0.10, 0.43) | 0.28 (0.13, 0.50) | 0.28 (0.12, 0.50) | 0.27 (0.13, 0.49) | 0.28 (0.14, 0.50) | 0.28 (0.13, 0.50) | 0.28 (0.13, 0.51) | 0.27 (0.12, 0.50) |
| Zip Code - PTs per 1000 | 0.16 (0.04, 0.37) | 0.19 (0.05, 0.44) | 0.20 (0.05, 0.45) | 0.20 (0.06, 0.46) | 0.20 (0.06, 0.47) | 0.20 (0.06, 0.47) | 0.20 (0.06, 0.46) | 0.19 (0.05, 0.45) |
| Zip Code - LAcs per 1000 | 0.00 (0.00, 0.03) | 0.00 (0.00, 0.03) | 0.00 (0.00, 0.03) | 0.00 (0.00, 0.03) | 0.00 (0.00, 0.03) | 0.00 (0.00, 0.03) | 0.00 (0.00, 0.03) | 0.00 (0.00, 0.03) |
| Clean Period Before Episode - days | 713 (478, 902) | 733 (482, 916) | 681 (461, 873) | 622 (431, 842) | 579 (409, 821) | 544 (373, 774) | 515 (310, 730) | 671 (459, 879) |
| Clean Period After Episode - days | 413 (239, 650) | 414 (237, 659) | 403 (232, 629) | 392 (230, 599) | 384 (232, 560) | 378 (229, 524) | 375 (230, 508) | 397 (235, 627) |
| **Initial Contact With Physical Therapist (PT) - # of Visits of Active Care (AC)** | | | | | | | | |
| # of PTs | 91 | 1038 | 738 | 485 | 342 | 231 | 590 | 2711 |
| % of PTs | 3.4% | 38.3% | 27.2% | 17.9% | 12.6% | 8.5% | 21.8% | 100.0% |
| Episodes/Individuals | 160 | 1319 | 893 | 568 | 404 | 260 | 844 | 4448 |
| % of Episodes/Individuals | 3.6% | 29.7% | 20.1% | 12.8% | 9.1% | 5.8% | 19.0% | 100.0% |
| Individuals - % Female | 50.6% | 57.2% | 60.6% | 58.5% | 58.9% | 59.6% | 60.9% | 58.8% |
| Individuals - Age | 46 (36, 56) | 45 (34, 55) | 45 (34, 55) | 46 (35, 56) | 44 (34, 54) | 47 (35, 56) | 45 (34, 55) | 45 (34, 55) |
| Individuals - ERG® Risk Score | 1.9 (0.8, 3.7) | 1.5 (0.8, 3.1) | 1.6 (0.8, 3.3) | 1.7 (0.9, 3.4) | 1.7 (0.8, 3.3) | 1.7 (0.8, 3.6) | 1.9 (0.9, 3.5) | 1.7 (0.8, 3.4) |
| Zip Code - % non-Hispanic White | 76% (59, 86) | 74% (54, 85) | 75% (54, 84) | 74% (57, 85) | 73% (58, 83) | 71% (55, 83) | 72% (53, 83) | 73% (55, 84) |
| Zip Code - Area Deprivation Index | 41 (24, 54) | 36 (22, 54) | 34 (21, 52) | 33 (19, 51) | 32 (20, 51) | 32 (19, 53) | 28 (15, 47) | 34 (19, 52) |
| Zip Code - DCs per 1000 | 0.24 (0.11, 0.46) | 0.27 (0.12, 0.49) | 0.27 (0.12, 0.46) | 0.27 (0.12, 0.51) | 0.28 (0.14, 0.49) | 0.28 (0.09, 0.47) | 0.26 (0.12, 0.48) | 0.27 (0.12, 0.48) |
| Zip Code - PTs per 1000 | 0.26 (0.06, 0.59) | 0.24 (0.08, 0.58) | 0.23 (0.08, 0.56) | 0.26 (0.09, 0.58) | 0.27 (0.11, 0.56) | 0.25 (0.08, 0.59) | 0.28 (0.10, 0.63) | 0.25 (0.09, 0.58) |
| Zip Code - LAcs per 1000 | 0.00 (0.00, 0.05) | 0.00 (0.00, 0.05) | 0.00 (0.00, 0.05) | 0.00 (0.00, 0.06) | 0.00 (0.00, 0.06) | 0.00 (0.00, 0.06) | 0.00 (0.00, 0.06) | 0.00 (0.00, 0.06) |
| Clean Period Before Episode - days | 626 (459, 856) | 689 (458, 879) | 646 (436, 851) | 614 (442, 824) | 620 (458, 824) | 633 (416, 843) | 558 (380, 790) | 628 (444, 842) |
| Clean Period After Episode - days | 406 (224, 615) | 400 (236, 652) | 411 (242, 657) | 400 (258, 628) | 428 (273, 596) | 396 (246, 645) | 402 (278, 626) | 406 (248, 643) |
| **Initial Contact With Physical Therapist (PT) - # of Visits of Manual Therapy (MT)** | | | | | | | | |
| # of PTs | 638 | 1067 | 655 | 381 | 243 | 129 | 168 | 2711 |
| % of PTs | 23.5% | 39.4% | 24.2% | 14.1% | 9.0% | 4.8% | 6.2% | 100.0% |
| Episodes/Individuals | 1144 | 1427 | 817 | 447 | 281 | 138 | 194 | 4448 |
| % of Episodes/Individuals | 25.7% | 32.1% | 18.4% | 10.0% | 6.3% | 3.1% | 4.4% | 100.0% |
| Individuals - % Female | 55.1% | 57.1% | 62.2% | 63.5% | 61.6% | 58.0% | 64.9% | 58.8% |
| Individuals - Age | 46 (36, 56) | 44 (34, 54) | 45 (34, 54) | 46 (35, 56) | 45 (33, 54) | 45 (35, 55) | 45 (36, 56) | 45 (34, 55) |
| Individuals - ERG® Risk Score | 1.7 (0.8, 3.5) | 1.6 (0.8, 3.2) | 1.6 (0.8, 3.3) | 1.7 (0.9, 3.2) | 1.5 (0.7, 3.3) | 2.0 (1.0, 4.0) | 2.0 (1.2, 3.4) | 1.7 (0.8, 3.4) |
| Zip Code - % non-Hispanic White | 74% (55, 85) | 73% (54, 83) | 73% (57, 84) | 73% (55, 83) | 74% (59, 84) | 71% (52, 85) | 72% (53, 84) | 73% (55, 84) |
| Zip Code - Area Deprivation Index | 37 (23, 55) | 34 (19, 52) | 31 (19, 50) | 32 (19, 50) | 31 (15, 52) | 29 (17, 49) | 24 (13, 43) | 34 (19, 52) |
| Zip Code - DCs per 1000 | 0.27 (0.11, 0.49) | 0.26 (0.11, 0.48) | 0.27 (0.13, 0.46) | 0.27 (0.13, 0.47) | 0.29 (0.15, 0.49) | 0.26 (0.11, 0.44) | 0.31 (0.12, 0.53) | 0.27 (0.12, 0.48) |
| Zip Code - PTs per 1000 | 0.22 (0.06, 0.51) | 0.26 (0.08, 0.60) | 0.26 (0.11, 0.58) | 0.27 (0.10, 0.65) | 0.28 (0.11, 0.62) | 0.25 (0.11, 0.57) | 0.36 (0.14, 0.67) | 0.25 (0.09, 0.58) |
| Zip Code - LAcs per 1000 | 0.00 (0.00, 0.04) | 0.00 (0.00, 0.05) | 0.00 (0.00, 0.07) | 0.00 (0.00, 0.06) | 0.00 (0.00, 0.07) | 0.00 (0.00, 0.04) | 0.02 (0.00, 0.08) | 0.00 (0.00, 0.06) |
| Clean Period Before Episode - days | 662 (458, 860) | 668 (457, 864) | 633 (436, 842) | 605 (415, 826) | 565 (387, 787) | 540 (333, 773) | 480 (272, 632) | 628 (444, 842) |
| Clean Period After Episode - days | 391 (225, 621) | 417 (245, 656) | 417 (256, 657) | 432 (265, 661) | 407 (294, 603) | 400 (290, 645) | 397 (354, 576) | 406 (248, 643) |
| **Initial Contact with Licensed Acupuncturist (LAc) - # of Visits of Acupuncture (Acu)** | | | | | | | | |
| # of LAcs | 64 | 502 | 504 | 204 | 205 | 85 | 272 | 1326 |
| % of LAcs | 4.8% | 37.9% | 38.0% | 15.4% | 15.5% | 6.4% | 20.5% | 100.0% |
| Episodes/Individuals | 233 | 644 | 622 | 228 | 233 | 90 | 339 | 2389 |
| % of Episodes/Individuals | 9.8% | 27.0% | 26.0% | 9.5% | 9.8% | 3.8% | 14.2% | 100.0% |
| Individuals - % Female | 68.7% | 64.1% | 63.0% | 68.9% | 67.4% | 58.9% | 68.7% | 65.5% |
| Individuals - Age | 41 (33, 50) | 38 (32, 49) | 40 (33, 49) | 40 (32, 50) | 41 (33, 52) | 41 (34, 50) | 43 (35, 51) | 40 (33, 50) |
| Individuals - ERG® Risk Score | 1.5 (0.6, 3.0) | 1.0 (0.4, 2.4) | 0.9 (0.4, 2.1) | 1.1 (0.5, 2.0) | 1.0 (0.4, 2.4) | 1.1 (0.5, 1.9) | 1.3 (0.5, 2.6) | 1.0 (0.4, 2.4) |
| Zip Code - % non-Hispanic White | 55% (37, 69) | 62% (40, 76) | 64% (43, 78) | 63% (47, 79) | 62% (41, 78) | 64% (37, 78) | 53% (34, 71) | 61% (40, 76) |
| Zip Code - Area Deprivation Index | 29 (19, 47) | 24 (14, 39) | 22 (12, 35) | 20 (10, 32) | 22 (12, 34) | 21 (9, 30) | 19 (11, 34) | 22 (12, 36) |
| Zip Code - DCs per 1000 | 0.27 (0.16, 0.51) | 0.25 (0.11, 0.48) | 0.27 (0.13, 0.51) | 0.34 (0.15, 0.56) | 0.28 (0.14, 0.54) | 0.34 (0.16, 0.52) | 0.24 (0.11, 0.49) | 0.27 (0.13, 0.51) |
| Zip Code - PTs per 1000 | 0.21 (0.07, 0.47) | 0.24 (0.09, 0.55) | 0.26 (0.10, 0.57) | 0.34 (0.13, 0.72) | 0.22 (0.09, 0.55) | 0.26 (0.15, 0.61) | 0.21 (0.07, 0.50) | 0.25 (0.09, 0.57) |
| Zip Code - LAcs per 1000 | 0.04 (0.00, 0.16) | 0.06 (0.00, 0.19) | 0.06 (0.00, 0.21) | 0.08 (0.02, 0.23) | 0.07 (0.00, 0.22) | 0.10 (0.00, 0.27) | 0.07 (0.01, 0.18) | 0.06 (0.00, 0.20) |
| Clean Period Before Episode - days | 681 (465, 858) | 708 (463, 915) | 715 (480, 913) | 718 (480, 903) | 651 (458, 891) | 580 (349, 785) | 579 (396, 830) | 681 (458, 891) |
| Clean Period After Episode - days | 440 (278, 652) | 430 (238, 676) | 406 (231, 640) | 387 (203, 648) | 448 (230, 659) | 470 (298, 698) | 391 (228, 586) | 416 (234, 652) |

Cells with red text denote that the effect of provider type on service usage was found not to be significantly different from that of PCP-reference (Mann-Whitney U $p > 0.001$)
Cells with black text denote that the effect of provider type on service usage was found to be significantly different from that of PCP-reference (Mann-Whitney U $p < 0.001$)
